# Updating the epidemiology of blastomycosis and histoplasmosis in the United States, using national electronic health record data, 2013 - 2023

**DOI:** 10.1101/2025.06.03.25328884

**Authors:** Juliana G.E. Bartels, Simon K. Camponuri, Theo T. Snow, Brittany L. Morgan Bustamante, Natalie J. Kane, Rose M. Reynolds, Aidan Lee, Mark Hoffman, Theodore C. White, Justin V. Remais, Jennifer R. Head

## Abstract

**Introduction:** Where surveillance data are limited, nationally-representative electronic health records allow for geographic, temporal, and demographic characterization of the fungal diseases blastomycosis and histoplasmosis.

**Methods:** We identified incident blastomycosis and histoplasmosis cases from 2013 to 2023 within Oracle EHR Real-World Data, which comprises 1.6 billion healthcare encounters nationally. To characterize geographic and temporal incidence rates, we used weighted generalized estimating equations adjusting for non-representativeness of EHR-reporting facilities. We computed standardized incidence rate ratios (sIRRs), which relay relative differences in standardized incidence rates among region, race/ethnicity, gender, and age subgroups and the national population.

**Results:** National incidence rates in 2023 were 2.4 (95% CI: 1.6-3.5) and 1.9 times (95% CI: 1.6-2.2) rates in 2013, for blastomycosis and histoplasmosis, respectively. Blastomycosis incidence rates among Hispanic/Latino and non-Hispanic Black individuals were 60% (sIRR: 1.6 [95% CI: 1.0-2.4]) and 30% (sIRR: 1.3 [95% CI: 1.0-1.6]) higher than the standardized national incidence rate. Histoplasmosis incidence rates were elevated among non-Hispanic White patients (sIRR: 1.05 [95% CI: 1.02-1.08]). Standardized incidence rates of both diseases were higher among older and male patients and were elevated in the Upper Midwest (sIRR: blastomycosis: 5.1 [95% CI: 3.7-6.8]; histoplasmosis: 1.7 [95% CI: 1.5-1.9]) and Ohio Valley regions (sIRR: blastomycosis: 2.0 [95% CI: 1.7-2.3]; histoplasmosis: 2.3 [95% CI: 2.2-2.5], and increased in the Northern Rockies and Plains from 2013 to 2023.

**Discussion:** This analysis revealed increasing incidence rates of blastomycosis and histoplasmosis and expansion outside of historically endemic regions, with notable differences in incidence by race/ethnicity, gender, and age.

**Summary:** Blastomycosis and histoplasmosis incidence rates have increased two-fold in the United States from 2013 to 2023, including outside of historically endemic regions. Incidence rates varied by race and ethnicity, age, and gender.

## Introduction

Blastomycosis and histoplasmosis are fungal diseases caused by infection with dimorphic fungal pathogens that persist in the environment.^1,2^ Case reports of blastomycosis and histoplasmosis indicate a shift outside of their historical geographic endemic range,^3–5^ potentially due to increasing use of immunosuppressive medications, migration from or travel to endemic areas,^6^ and climate change.^7–9^ In the United States, blastomycosis and histoplasmosis are reportable in only 8 and 15 states, respectively, limiting our understandings of their geographic range. Additionally, missing demographic information within these surveillance datasets has limited our understanding of differences in incidence by gender, race, and ethnicity.^2,10^

Blastomycosis is caused by inhalation of *Blastomyces dermatitidis* and *B. gilchristii*, which occupy moist, acidic soil and rotting wood.^11^ In the U.S., *Blastomyces spp.* are considered endemic in the Great Lakes region, several states in the Mississippi and Ohio River valleys, and along the St. Lawrence River in New York and Canada.^11^ Histoplasmosis, caused by inhalation of spores of *Histoplasma capsulatum* found in soil, is endemic in the Mississippi and Ohio River valleys, although its distribution is likely national.^12,13^ Most blastomycosis and histoplamosis infections are sporadically-acquired from environmental exposures;^12^ point-source outbreaks have been linked to soil excavation,^14^ yard waste,^15^ paper milling,^16^ outdoor recreation,^17^ and, for histoplasmosis, disturbance of bird or bat droppings.^1^ Blastomycosis and histoplasmosis are often underdiagnosed due to limitations in diagnostics,^18^ non-specific clinical presentation similar to community-acquired bacterial pneumonia,^19^ and varying degrees of diagnostic suspicion.^20–22^

Public health leaders have called for epidemiologic studies to understand the distribution and geographic range of blastomycosis and histoplasmosis, and have advised using longitudinal electronic health record (EHR) data to complement public health surveillance systems.^23,24^ Preliminary assessments using EHR data have advanced our understanding of fungal disease distribution in the U.S., but have been limited to local health systems or inpatient populations.^25,26^ Here, we leveraged data from Oracle EHR Real-World Data (OERWD)—an EHR database with national coverage—to conduct a retrospective cohort study examining the geographic and temporal distribution of blastomycosis and histoplasmosis incidence in the U.S. from 2013 to 2023, and quantify differences in incidence rates by race and ethnicity, gender, and age.

## Methods

### Data Source

OERWD comprises de-identified electronic medical records of hospitals that have a data use agreement with Oracle, in compliance with Health Insurance Portability and Accountability Act requirements. OERWD contains encounter information for over 109 million patients and over 1.6 billion healthcare encounters over two decades. Encounters may include pharmacy, laboratory, admission, and billing information which are date and time stamped.

### Ethics

De-identified fungal disease data derived from OERWD were provided through a Data Transfer and Use Agreement with Children’s Mercy Hospital, Kansas City, Missouri. The Children’s Mercy Office of Research Integrity deems work with OERWD to be non-human subjects research.

### Case Definition and Estimation of Person-Time At-Risk

Patients with at least one inpatient or outpatient encounter between January 1, 2013 and December 31, 2023 were eligible for inclusion; patients missing discharge dates or state of residence were excluded. Patients were categorized into U.S. census racial and ethnic categories using self-reported race and ethnicity provided by Oracle (see Supplemental Methods),^27^ five-year age groups using birthdate and encounter date, and National Oceanic and Atmospheric Association climate regions using state location (excluding Alaska and Hawaii).^28^ We identified incident case-patients using their initial encounter with an International Classification of Diseases (ICD)-9 or ICD-10 code for blastomycosis (ICD-9: 116.0; ICD-10: B40*) or histoplasmosis (ICD-9: 115*; ICD-10: B39*) in any position of the record; case-patients whose first diagnosis occurred before 2013 were excluded. Patients contributed person-time (i.e., time at risk for disease) for years in which they had a diagnosis for any condition. We used post-stratification weights to adjust for differences in age, gender, race and ethnicity, state, and year between our sample and the U.S. population resulting from non-representativeness of facilities that report to OERWD (see Supplemental Methods).

### Geographic and temporal distribution of blastomycosis and histoplasmosis incidence

We calculated annual incidence rates within climate regions by fitting region-specific generalized estimating equation (GEE) Poisson regression models with post-stratification weights to case data, with fixed effects on year (*y*), and an offset for the log of weighted patient-years, as log(*E*[*Y*|***β***, *y*]) = ***β*** ∗ *y* + log (*patient* − *years*). To examine how incidence rates changed within states over time, we stratified the data by four time periods (2013-2015, 2016-2018, 2019-2021, 2023-2023) and fit period-specific weighted GEE Poisson models with fixed effects for state (*s*), as log(*E*[*Y*|***β***, *s*]) = ***β*** ∗ *s* + log (*patient* − *years*). To estimate state-, age-, gender-, and race and ethnicity-adjusted incidence rate ratios (aIRRs) reflecting the change in incidence rates over time for the overall U.S., we used weighted GEE Poisson models, as log(*E*[*Y*|*β*, ***X***]) = ***Xβ*** + log (*patient* − *years*), where ***X*** represents a matrix of covariates including fixed effects for year, state, age group, gender, and racial and ethnic group.

We used direct standardization to compute incidence rates that would have been observed in each region if the distribution of age, gender, and race and ethnicity matched the U.S. population (our reference population; see Supplemental Methods), enabling comparison of incidence rates across regions. Standardized incidence rates (sIRs) by age group, racial and ethnic group, and gender were also calculated using direct standardization (see Supplemental Methods). We generated 1000 cluster bootstrapped datasets from the original dataset stratified by state-month clusters, and calculated 95% confidence intervals (CIs) as the 2.5^th^ and 97.5^th^ percentile of the distribution using the R *boot* package.^29^

### Differences in incidence by geography and demographics

To evaluate whether regions, age groups, racial and ethnic groups, and genders had disproportionately high blastomycosis or histoplasmosis sIRs, we computed overall and year-specific standardized incidence rate ratios (sIRRs) comparing the group-specific sIR to the sIR in the standard (national) population (see Supplemental Methods). An sIRR greater than 1 indicates a region or group had a higher incidence rate than the national sIR, accounting for varying distributions of demographic characteristics in the population; an sIRR less than 1 indicates lower incidence rate than the national sIR.

Estimating differences using sIRRs rather than incidence rate ratios from regression models allows us to compare group-specific incidence rates to the national average rather than selecting one group to serve as the reference. In the case of racial differences, for instance, this allows us to de-center one socially-constructed racial and ethnic group as the reference population.^30^ All analyses were completed in R 4.4.1, using *geepack* for GEE Poisson regressions.^31^

## Results

### Study population

Our cohort included 8,313 histoplasmosis case-patients, 1,424 blastomycosis case-patients, and 168,316,366 patient-years from 2013 to 2023, after excluding 19 blastomycosis cases, 57 histoplasmosis cases, and 3,274,376 patient-years missing state location (**Table 1**). Female patients contributed more patient-years (55.9%) and slightly more histoplasmosis case-patients (51.7%), while the majority of blastomycosis case-patients were male (69.3%; Table 1). The majority of case-patients were non-Hispanic (NH) White (blastomycosis: 73.0%; histoplasmosis: 84.1%). NH White individuals also contributed the most patient-years (62.8%). Older age groups made up the majority of case-patients (blastomycosis: 55 and older, 62.1%; histoplasmosis: 55 and older, 58.8%), and most case-patients resided in the Ohio Valley (blastomycosis: 40.5%; histoplasmosis: 52.7%; **Table 1**).

**Table 1.** Number and proportion of blastomycosis and histoplasmosis cases and total Oracle Cerner Real-World Data (OERWD) patient-years, by patient characteristics – United States, 2013-2023.

| Characteristic | Blastomycosis<br>N (%) | Histoplasmosis<br>N (%) | OERWD patient-years<br>N (%) |
| --- | --- | --- | --- |
| N | 1,422 (100.0) | 8,313 (100.0) | 168,316,366 (100.0) |
| Gender |  |  |  |
| Female | 437 (30.7) | 4,295 (51.7) | 94,024,336 (55.9) |
| Male | 985 (69.3) | 4,018 (48.3) | 74,223,588 (44.1) |
| Age Group |  |  |  |
| Under 5 | 10 (0.7) | 45 (0.5) | 15,257,181 (9.1) |
| 5 to 9 | 26 (1.8) | 97 (1.2) | 10,832,218 (6.4) |
| 10 to 14 | 26 (1.8) | 157 (1.9) | 10,309,671 (6.1) |
| 15 to 19 | 54 (3.8) | 312 (3.8) | 10,876,186 (6.5) |
| 20 to 24 | 30 (2.1) | 193 (2.3) | 9,225,530 (5.5) |
| 25 to 29 | 45 (3.2) | 230 (2.8) | 8,933,086 (5.3) |
| 30 to 34 | 63 (4.4) | 315 (3.8) | 9,232,681 (5.5) |
| 35 to 39 | 53 (3.7) | 387 (3.8) | 8,890,755 (5.3) |
| 40 to 44 | 62 (4.4) | 448 (5.4) | 8,737,494 (5.2) |
| 45 to 49 | 82 (5.8) | 576 (6.9) | 8,983,560 (5.3) |
| 50 to 54 | 89 (6.3) | 665 (8.0) | 10,081,815 (6) |
| 55 to 59 | 129 (9.1) | 793 (9.5) | 10,893,956 (6.5) |
| 60 to 64 | 156 (11.0) | 897 (10.8) | 11,051,869 (6.6) |
| 65 and over | 597 (42.0) | 3,198 (38.5) | 35,001,125 (20.8) |
| Race and Ethnicity<sup>a</sup> |  |  |  |
| Hispanic or Latino | 154 (11.1) | 513 (6.3) | 28,720,842 (17.7) |
| NH AI/AN | 10 (0.7) | 15 (0.2) | 1,240,457 (0.8) |
| NH Asian | 42 (3.0) | 86 (1.1) | 4,335,822 (2.7) |
| NH Black or African American | 136 (9.8) | 529 (6.5) | 17,696,116 (10.9) |
| NH Other Race | 32 (2.3) | 150 (1.8) | 8,469,405 (5.2) |
| NH White | 1010 (73.0) | 6,864 (84.1) | 102,202,122 (62.8) |
| Climate Region<sup>b</sup> |  |  |  |
| Northeast | 173 (12.2) | 582 (7.0) | 41,308,306 (24.6) |
| Northern Rockies and Plains | 35 (2.5) | 561 (6.8) | 6,199,851 (3.7) |
| Northwest | 10 (0.7) | 38 (0.5) | 2,383,155 (1.4) |
| Ohio Valley | 574 (40.5) | 4,380 (52.7) | 35,686,542 (21.3) |
| South | 48 (3.4) | 780 (9.4) | 11,430,832 (6.8) |
| Southeast | 54 (3.8) | 352 (4.2) | 21,656,428 (12.9) |
| Southwest | 51 (3.6) | 407 (4.9) | 18,312,052 (10.9) |
| Upper Midwest | 261 (18.3) | 778 (9.4) | 7,329,988 (4.4) |
| West | 213 (15.0) | 432 (5.2) | 23,431,004 (14) |

Abbreviations: NH = non-Hispanic, AI/AN = American Indian or Alaska Native.
<sup>a</sup> Race and ethnicity were unknown for 38 patients (2.7%) with a blastomycosis diagnosis, 156 patients (1.9%) with a histoplasmosis diagnosis, and 6,892,059 OERWD patient-years (4.1%).
<sup>b</sup> Climate regions do not include Alaska and Hawaii. 3 patients (0.2%) with a blastomycosis diagnosis were diagnosed in Alaska and Hawaii, 3 patients (0.03%) with a histoplasmosis diagnosis were diagnosed in Alaska or Hawaii; and 578,208 OERWD patient-years were located in Alaska or Hawaii (0.3%).

### Spatiotemporal trends in incidence

#### Blastomycosis

The national incidence rate of blastomycosis per 100,000 patient-years (p-y) increased between 2013 and 2023 from 0.4 cases per 100,000 p-y (95% CI: 0.3-0.6) to 1.3 per 100,000 p-y (95% CI: 1.1-1.6; **Figure 1**, **Table S2**). The adjusted blastomycosis incidence rate in 2023 was 2.4 times (95% CI: 1.6-3.5) the rate in 2013 (**Figure 2**). Blastomycosis incidence increased in all regions over the study period (**Table S1**; **Table S2**), with the most profound increases in the Upper Midwest (0.5 cases per 100,000 p-y in 2013 to 6.8 cases per 100,000 p-y in 2023; **Figure 1**). Most states maintained incidence rates between 0-2.0 cases per 100,000 p-y from 2013-2023. However, incidence rates exceeded 4.0 cases per 100,000 p-y in 2022-2023 in some states in the Upper Midwest (e.g., Wisconsin: 9.9; Michigan: 6.2), the Ohio Valley, (e.g., Illinois: 4.1; West Virginia: 4.9), and the South (e.g., Arkansas: 10.7) (**Figure 1**, **Table S1**).

**Figure 1.**
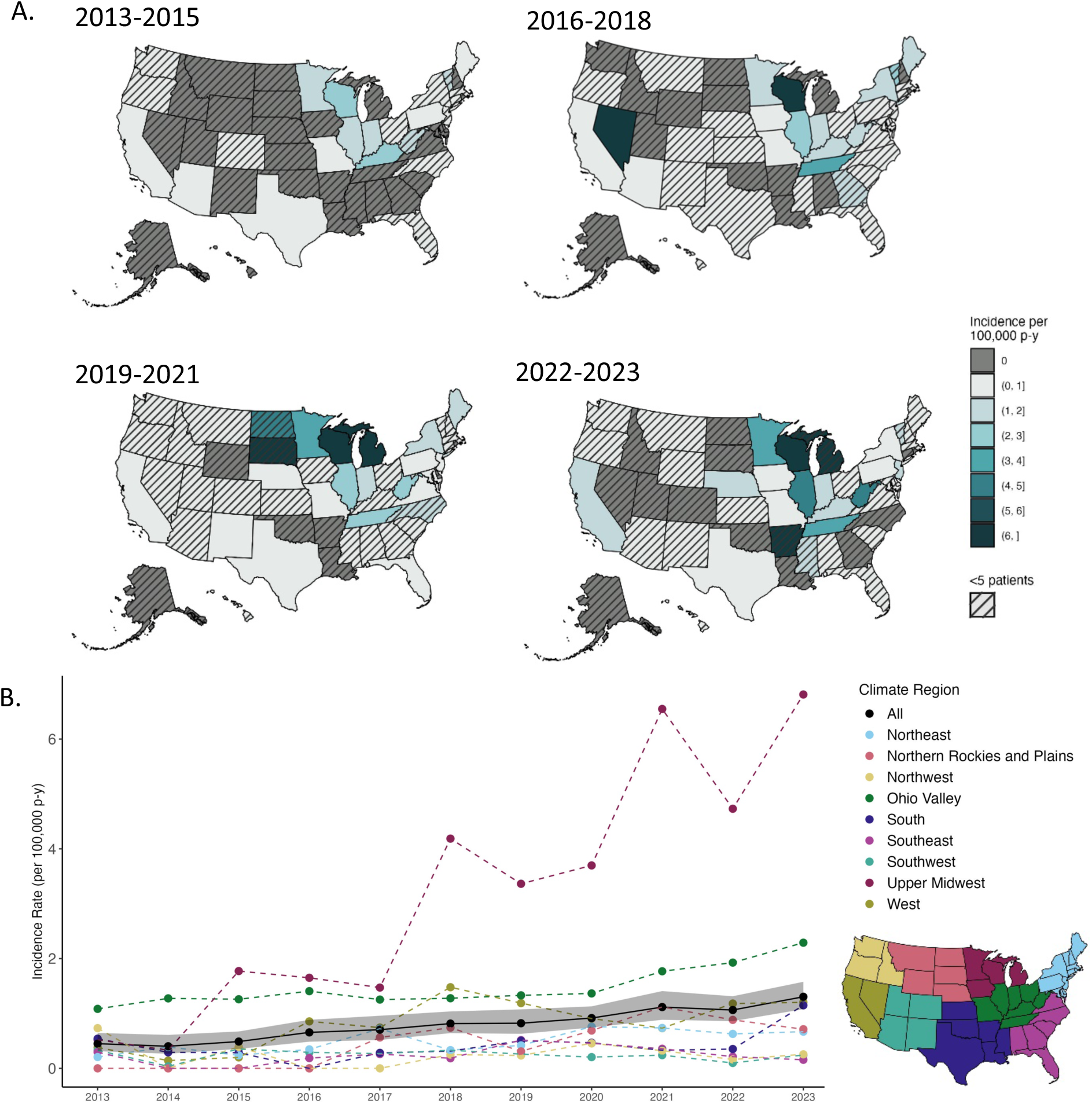
Incidence rate of blastomycosis per 100,000 patient-years by state and climate region, 2013-2023. A. Incidence rates of blastomycosis by state and period: 2013-2015, 2016-2018, 2019-2021, and 2022-2023. Hatches indicate states that had less than 5 blastomycosis cases during the time period. B. Incidence rate of blastomycosis per 100,000 patient-years by year and climate region (colored, dashed lines and points). Black points show the national annual incidence rate estimate, and the gray ribbon indicates the 95% CI. Inset map shows climate regions, which do not include Alaska or Hawaii. See Tables S1 and S2. Alt text: Four maps showing incidence rates of blastomycosis for four time periods in all 50 US states above a graph showing the incidence rates of blastomycosis by climate region and in the US overall annually from 2013 to 2023.

**Figure 2.**
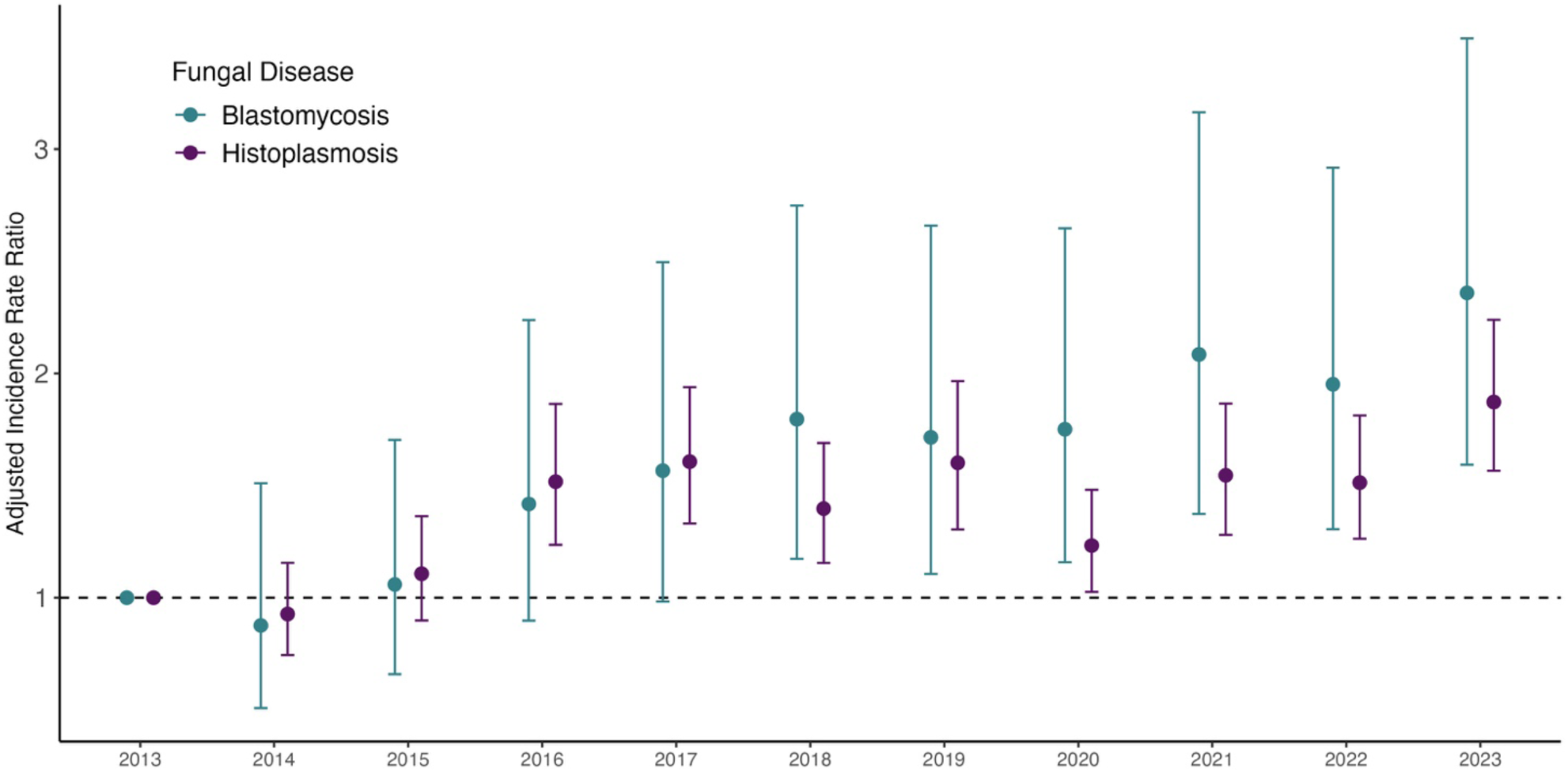
Annual adjusted incidence rate ratios for blastomycosis and histoplasmosis in the United States, 2013-2023. Annual adjusted incidence rate ratios (aIRRs) for blastomycosis and histoplasmosis in the United States compared to the reference year, 2013. Points indicate effect estimates and vertical bars show 95% CIs. aIRRs are adjusted for state, gender, race and ethnicity, and age group. Alt text: Graph showing annual adjusted incidence rate ratios of blastomycosis and histoplasmosis for 2013 to 2023 compared to the incidence rate in 2013 for each disease.

During the study period (2013-2023), the standardized incidence rates (sIRs) in the Upper Midwest and Ohio Valley were 5.1 and 2.0 times that of the national patient population (sIRR: Upper Midwest: 5.1 [95% CI: 3.7-6.8]; Ohio Valley: 2.0 [95% CI: 1.7-2.3]; **Figure 3A**; **Table S3**). The highest increases in sIR occurred in the Upper Midwest, from 0.4 cases per 100,000 p-y (95% CI: 0.0-0.9) in 2013 to 8.2 cases per 100,000 p-y (95% CI: 5.1-11.9) in 2023. In the Northern Rockies and Plains, the sIR increased from 0.4 cases per 100,000 p-y [95% CI: 0.0-0.9] in 2017 to 1.7 cases per 100,000 p-y (95% CI: 0.1-4.3) at the peak in 2022. The Southeast, South, Northeast, Southwest, and Northwest regions, had sIRs lower than the national average during the study period (sIRR: Southeast: 0.3 [95% CI: 0.2-0.4]; South: 0.5 [95% CI: 0.4-0.7]; Northeast: 0.6 [95% CI: 0.5-0.8]; Southwest: 0.3 [95% CI: 0.2-0.4]; Northwest: 0.2 [95% CI: 0.1-0.4] (**Figure 3A**; **Table S3**).

**Figure 3.**
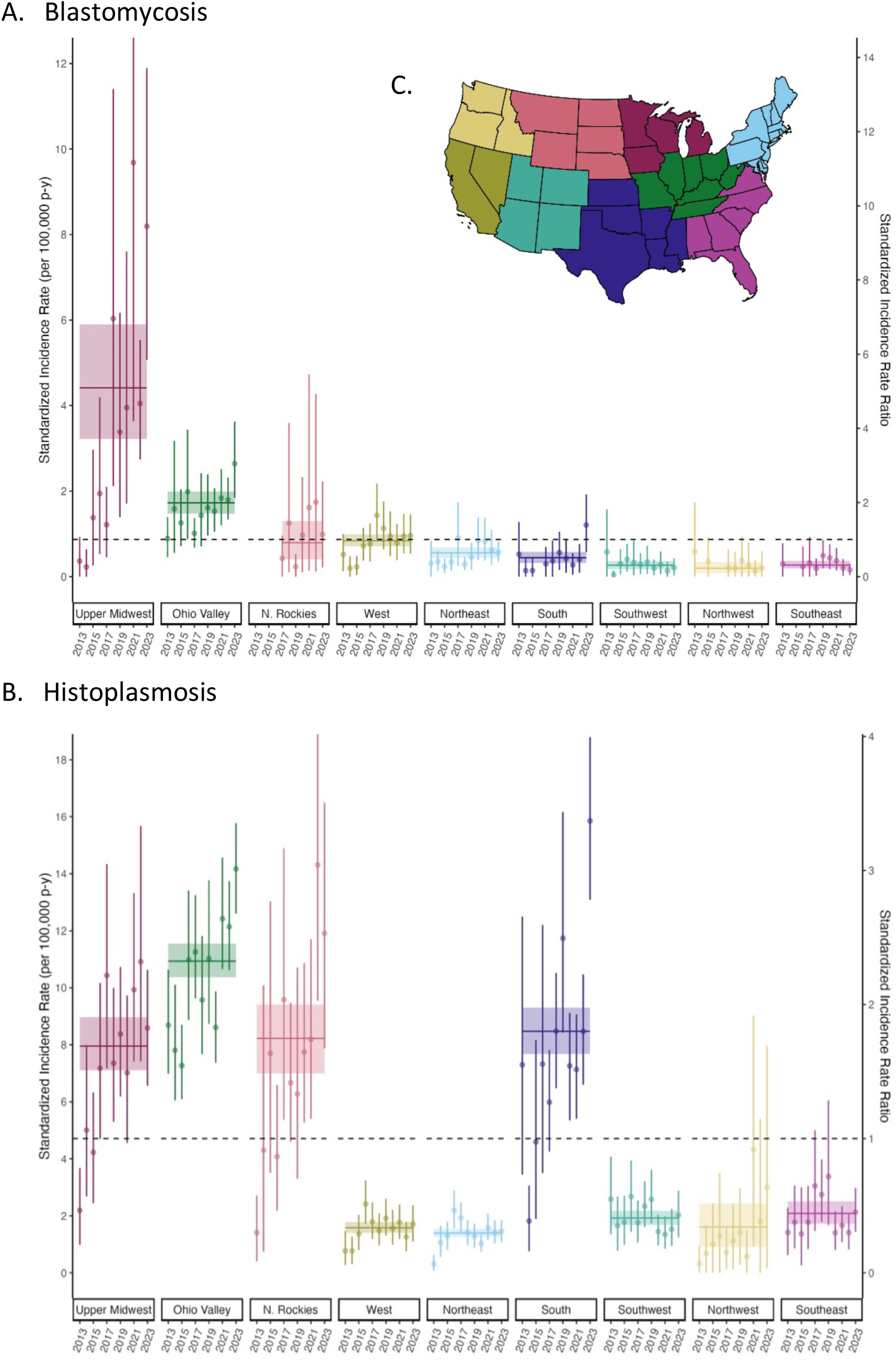
Standardized incidence rates and standardized incidence rate ratios of blastomycosis and histoplasmosis per 100,000 patient-years by climate region and year, United States, 2013-2023. A. Standardized incidence rate (sIR, left y-axis) and standardized incidence rate ratio (sIRR, right y-axis) of (A) blastomycosis and (B) histoplasmosis per 100,000 patient-years by climate region, 2013-2023. sIRs are direct standardized to the U.S. national population by gender, race and ethnicity, and age group. Points represent the sIR and sIRR for each year and vertical bars represent 95% CIs. The horizontal lines represent the sIR and sIRR for each climate region for the entire study period, and the colored boxes represent the 95% CIs for the entire study period. The dashed horizontal line shows the null sIRR of 1. N. Rockies = Northern Rockies and Plains. (C) A map of climate regions in the United States. Climate regions do not include Alaska and Hawaii. See Table S3. Alt text: Two graphs showing the annual and mean standardized incidence rates and standardized incidence rate ratios of blastomycosis and histoplasmosis in each climate region in the United States.

#### Histoplasmosis

National histoplasmosis incidence increased from 2.9 cases per 100,000 p-y in 2013 (95% CI: 2.4-3.5) to 6.5 cases per 100,000 p-y in 2023 (95% CI: 5.7-7.5; **Figure 4B**). The adjusted incidence rate in 2023 was 1.9 times that in 2013 (95% CI: 1.6-2.2; **Figure 2**; **Table S2**). Incidence rates in most regions increased slightly over the study period, with the highest rates in most years in the Ohio Valley (**Figure 4B**, **Table S2**). Incidence rates increased each year in the Upper Midwest between 2013 and 2017 (2.8 cases per 100,000 p-y in 2013 to 11.8 cases per 100,000 p-y in 2017), and remained steady thereafter. Incidence rates in the Northern Rockies and Plains and the South surpassed the Upper Midwest in 2023 (Northern Rockies and Plains: 11.0 cases per 100,000 p-y; South: 13.3 per 100,000 p-y; Upper Midwest: 10.4 per 100,000 p-y). By 2022-2023, incidence rates increased to above 20 cases per 100,000 p-y in Indiana (21.0), Iowa (20.7), Kansas (28.6), and Nebraska (22.4), reflecting the increasing incidence in the Northern Rockies and Plains and South after 2017 (**Figure 4A**, **Table S2**).

**Figure 4.**
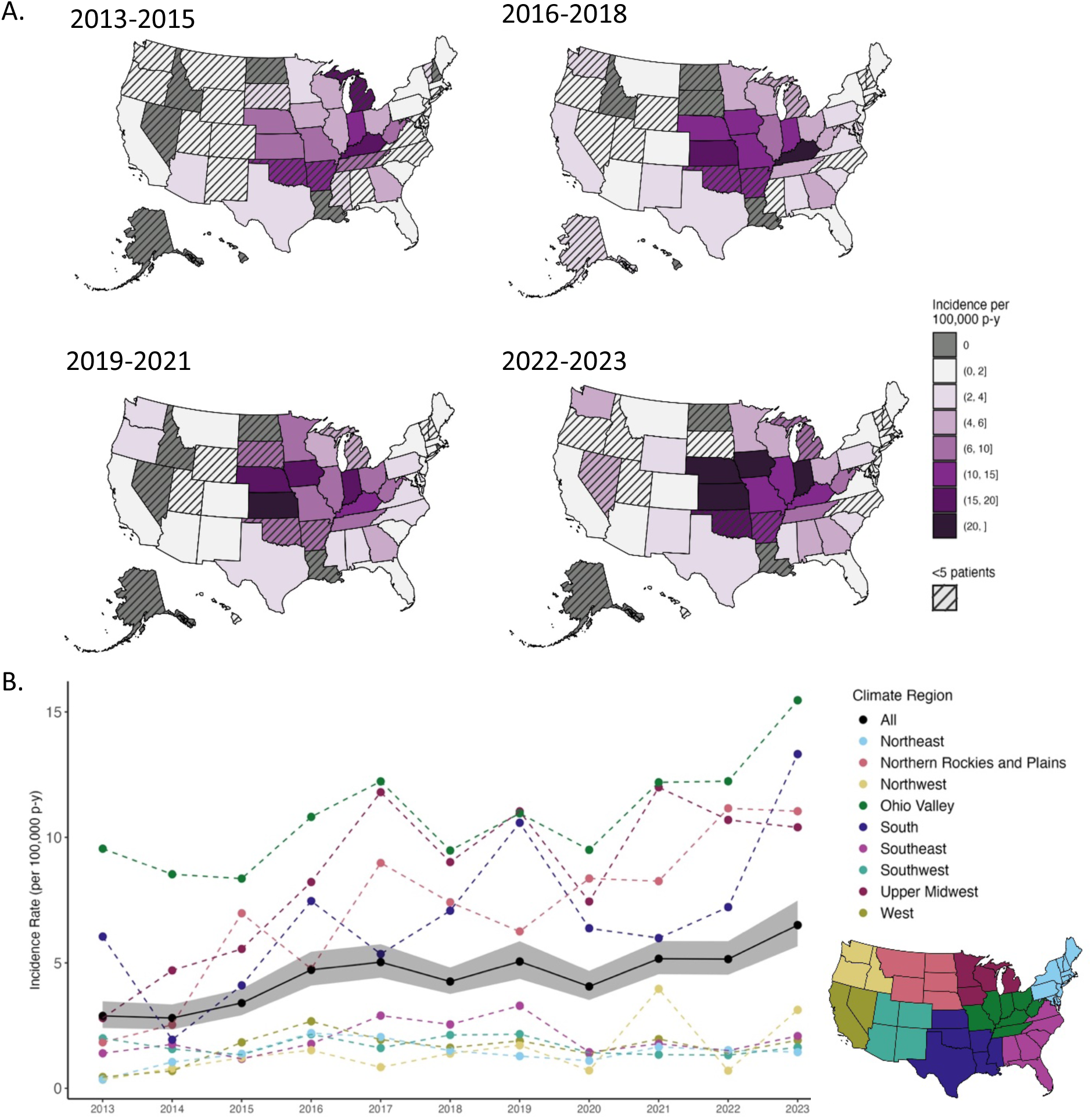
Incidence rate of histoplamosis per 100,000 patient-years by state and climate region, United States, 2013-2023. A. Incidence rate by state of histoplasmosis by period: 2013-2015, 2016-2018, 2019-2021, and 2022-2023. Hatches indicate states that had less than 5 histoplasmosis cases during the time period. B. Incidence rate of histoplasmosis per 100,000 patient-years by year and climate region (colored, dashed lines and points). Black points show national annual incidence rate estimate, and the gray ribbon shows 95% CI. Inset map shows climate regions, which do not include Alaska or Hawaii. See Tables S1 and S2. Alt text: Four maps showing incidence rates of histoplasmosis for four time periods in all 50 US states above a graph showing the incidence rates of histoplasmosis by climate region and in the US overall annually from 2013 to 2023.

The Upper Midwest and Ohio Valley had the highest mean sIRs from 2013-2023 (sIR: Upper Midwest: 8.0 cases per 100,000 p-y [95% CI: 7.1-9.0]; Ohio Valley: 10.9 cases per 100,000 p-[95% CI: 10.4-11.6]). The Northern Rockies and Plains sIR increased the most over the study period, from 1.4 cases per 100,000 p-y (95% CI: 0.4-2.7) in 2013 to the peak in 2022 of 14.3 cases per 100,000 p-y (95% CI: 9.5-20.0) (**Figure 4B**, **Table S3**). The Upper Midwest, Ohio Valley, Northern Rockies and Plains, and South all had disproportionately high sIRs compared to the national sIR (sIRR: Upper Midwest: 1.7 [95% CI: 1.5-1.9]; Ohio Valley: 2.3 [95% CI: 2.2-2.5]; Northern Rockies and Plains: 1.7 [95% CI: 1.5-2.0]; South: 1.8 [95% CI: 1.6-2.0]; **Figure 3B**; **Table S3**).

### Patient characteristics

The sIR of blastomycosis increased for each successive age group, from 0.1 cases per 100,000 p-y (95% CI: 0.0-0.1) among patients under 5 years to 1.9 cases per 100,000 p-y (95% CI: 1.7-2.1) for patients aged 65 and over (**Figure 5A**; **Table S4**). Patients older than 55 had higher sIRs compared to the national sIR for the study period (sIRR: 55-59: 1.6 [95% CI: 1.2-2.0]; 60-64: 1.8 [95% CI: 1.5-2.1]; 65+ (2.2 [95% CI: 1.9-2.4]). The sIRs for histoplasmosis also increased from 0.3 cases per 100,000 p-y (95% CI: 0.2-0.4) in patients under 5 to 8.4 cases per 100,000 p-y (95% CI: 7.9-8.9) among patients aged 65 and over. Patients older than 45 had disproportionately high histoplasmosis incidence compared to the national sIR (sIRR > 1) (**Figure 5B**; **Table S4**).

**Figure 5.**
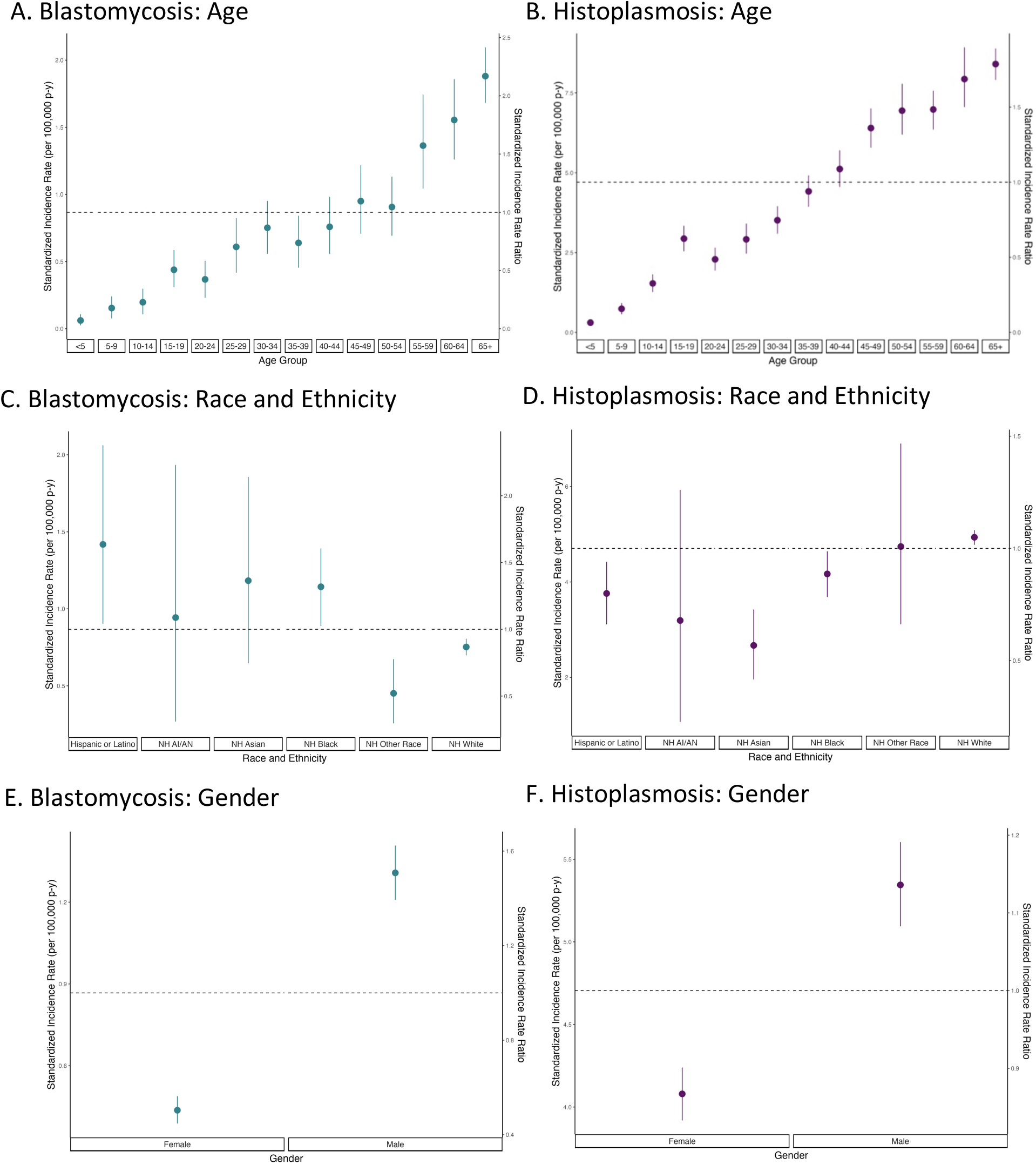
Standardized incidence rate and standardized incidence rate ratio of blastomycosis and histoplasmosis per 100,000 patient-years by age group, race and ethnicity, and gender, United States, 2013-2023. Standardized incidence rate (sIR, left y-axis) and standardized incidence rate ratio (sIRR, right y-axis) of blastomycosis (A, C, E) and histoplasmosis (B, D, F) per 100,000 patient-years by age group (A, B), race and ethnicity (C, D), and gender (E, F) 2013-2023. sIRs are direct standardized to the U.S. national population for gender, race and ethnicity, and state (A, B); gender, state, and age group (C, D); and state, race and ethnicity, and age group (E, F). Points represent the sIR and sIRR compared to the U.S. sIR for each patient characteristic; vertical lines represent 95% CIs. The dashed line shows the U.S. sIR for the total population and the null sIRR of 1. NH = non-Hispanic; AI/AN = American Indian or Alaska Native; NH Black = non-Hispanic Black or African American. See Table S4. Alt text: Six graphs showing mean standardized incidence rates and standardized incidence rate ratios of blastomycosis and histoplasmosis by age, gender, and race and ethnicity in the United States.

Hispanic or Latino patients and NH Black or African American patients had 60% and 30% higher sIRs of blastomycosis, respectively, over the study period compared to the national sIR (sIRR: Hispanic or Latino: 1.6 [95% CI: 1.0-2.4]; NH Black or African American: 1.3 [95% CI: 1.0-1.6]; **Figure 5C**; **Table S4**). NH AI/AN and NH Asian individuals also experienced elevated blastomycosis incidence rates compared to the national rate, although not significantly so, and the sample size was very small (sIRR: NH AI/AN: 1.1 [95% CI: 0.3–2.2]; NH Asian: 1.4 [95% CI: 0.7-2.1]). The only racial and ethnic group to experience a disproportionately high histoplasmosis incidence compared to their national representation was NH White (sIRR: 1.05 [95% CI: 1.02-1.08]; **Figure 5D**; **Table S4**). Hispanic and Latino, NH Asian, and NH Black or African American patients had disproportionately low histoplasmosis incidence rates during the study (sIRR: Hispanic or Latino: 0.8 [95% CI: 0.7-0.9]; NH Asian: 0.6 [95% CI: 0.4-0.7]; NH Black or African American: 0.9 [95% CI: 0.8-1.0]; **Figure 5D**).

Male patients had 50% higher blastomycosis and 10% higher histoplasmosis sIRs than the national sIR (sIRR: blastomycosis: 1.5 [95% CI: 1.4-1.6], histoplasmosis: 1.1 [95% CI: 1.1-1.2]; **Figure 5E, 5F**; **Table S4**). The sIR of blastomyosis for male patients (1.3 cases per 100,000 p-y [95% CI: 1.2-1.4]) was more than three times that of female patients (0.4 cases per 100,000 p-y [95% CI: 0.4-0.5]). The sIR for histoplasmosis among male patients was also greater than for female patients (sIR: male: 5.3 cases per 100,000 p-y [95% CI: 5.1-5.6]; female: 4.1 cases per 100,000 p-y [95% CI: 3.9-4.2]).

## Discussion

This retrospective cohort study using EHR data from 2013-2023 in the U.S. provides updated, nationally representative evidence that the geographic range of blastomycosis and histoplasmosis extends beyond historical endemic regions. Blastomycosis incidence increased 2.4 times from 2013 to 2023 and histoplasmosis incidence increased 1.9 times from 2013 to 2023. High incidence rates of both diseases occurred in states without surveillance data for these diseases, including Kansas and Nebraska.

We found that the blastomycosis and histoplasmosis diagnoses occurred beyond the historically endemic regions of the Mississippi and Ohio River valleys.^13,22^ While the Upper Midwest and Ohio Valley had the highest sIRs for histoplasmosis and blastomycosis from 2013-2023, the Northern Rockies and Plains and the South also had disproportionately high sIRs of histoplasmosis from 2013 to 2023, with high incidence in Kansas and Nebraska after 2017. Blastomycosis incidence in the Northern Rockies and Plains also increased steadily. States such as Nevada, Arkansas, and South Dakota had higher blastomycosis incidence in some time periods that subsequently subsided, which could indicate outbreaks or cases related to travel or migration. Other studies using data from major national commercial laboratory systems and Medicare insurance claims have also found that areas outside of the reportable geographies have increasing incidence of both diseases in the 21^st^ century,^22,32,33^ but the reasons for these changing geographic distributions remain to be thoroughly investigated.

Our study provides further evidence of incidence differences by racial and ethnic group, age group, and gender. We found that NH AI/AN, NH Asian, Hispanic or Latino patients, and NH Black or African American patients had higher sIRs of blastomycosis between 2013 and 2023. This aligns with surveillance data suggesting higher blastomycosis incidence rates among NH AI/AN and NH Asian populations,^10^ as well as documented outbreaks of blastomycosis among Indigenous and NH Asian populations in Wisconsin and Ontario, Canada.^3,34^ Several studies have found differences in clinical presentation, disease severity, and case fatality rates of blastomycosis among minority racial and ethnic patients compared to NH White patients, although understandings of structural and upstream causes were limited.^2,35,36^ Patients identifying as NH White experienced a disproportionately high sIR of histoplasmosis, while Hispanic and Latino, NH Asian, and NH Black or African American patients had disproportionately low histoplasmosis sIRs during the study period, similar to surveillance studies.^10^

Blastomycosis patients aged 55 and older and histoplasmosis patients aged 45 and older had the highest sIRs among age groups. This aligns with surveillance data showing most cases are among older adults.^10^ In addition, we found that the sIRs of both diseases were disproportionately high among male patients, with the sIR of blastomycosis among male patients more than three times that of female patients. These patterns are similar to gender ratios in surveillance and administrative data.^2^ Histoplasmosis surveillance data document more cases among male than female patients;^10^ although our cohort included slightly more female than male case-patients, the sIR was higher among males. Gender differences in blastomycosis incidence are often attributed to gendered exposure pathways (e.g., outdoor recreation,^17^ occupational exposure^16^), informing descriptions of a “typical” blastomycosis patient as a younger man who works or recreates outdoors.^37^ Further research is needed to examine whether observed gender differences are mediated by genderized exposure pathways,^37^ and to assess the possibility of underdiagnosis among female patients not matching the profile of a “typical” patient.

Future research should examine how factors like bat and bird migration, climatic changes in temperature and precipitation, travel, prevalence of immunosuppression, and diagnostic awareness influence the shifting geographic distributions of these diseases.^5,38^ Additionally, studies should investigate upstream drivers of racial and ethnic and gendered differences in hospitalization, mortality, and disease severity among blastomycosis and histoplasmosis patients, using additional data available in EHR, including prescriptions and labs.

### Limitations

EHR-based analyses are subject to selection bias due to systematic exclusion of patients who do not seek healthcare at data-contributing facilities; we used post-stratification weighting to make our sample more nationally representative and improve generalizability. Reported incidence rates likely underestimate true disease burdens due to missed diagnoses.

Overestimation relative to surveillance data is also possible, as patients only contributed person-time during healthcare encounters, whereas every resident in the state contributes person-time to surveillance denominators.

The study period includes years during which substantial changes to regulatory requirements for reporting and use of EHRs occured. Early EHR data may have been less complete or accurate, or exhibited other data capture issues that were mitigated as EHR technology and use improved over time. Case definitions relied on ICD codes, which may be only moderately sensitive for fungal disease cases.^39^ Incidence relied on the date of the initial encounter with an ICD code, rather than date of exposure or disease onset, and state locations recorded in OERWD refer to location of patient residence, but do not necessarily correspond to the location of patient exposure. Therefore, geographic estimates of disease burden represent spatial distribution of fungal patient residence rather than fungal exposures. Misclassification for cases who acquired infection during out-of-state travel, or who received a diagnosis later in life for an earlier asymptomatic infection after migration out-of-state is possible.^6,20^

## Data Availability

Data not publicly available.

## Acknowledgements

Data not publicly available.

JGEB and JRH jointly conceptualized the research study, outlining the main objectives and hypotheses. NJK, RMR, AL, and JGEB executed data processing, ensuring adherence to ethical standards and study protocols. JGEB led statistical analysis, with support and substantial contributions from SKC, TTS, JRH, and BLMB. JGEB and JRH carried out interpretation of study findings. JGEB wrote the original draft of the final manuscript, and all authors provided feedback. MH, TCW, and JVR provided guidance, access to the data, and supervision. JRH made the final decision to submit for publication.

## Supplementary Material

### Supplemental Methods

#### Categorization of patient race and ethnicity

Patients with Hispanic or Latino ethnicity were classified as Hispanic or Latino and patients with Non-Hispanic ethnicity were classified as non-Hispanic white (NH white), non-Hispanic Black or African American (NH Black or African American), non-Hispanic Asian (NH Asian), non-Hispanic American Indian or Alaska Native (NH AI/AN), and non-Hispanic Other Race (NH Other Race). Patients missing race data were classified “Unknown” if their ethnicity was missing or non-Hispanic.

#### Post-stratification weighting

Post-stratification weights were calculated by taking the ratio of the proportion of the national population comprising each demographic strata (age group, gender, and race and ethnicity) for each state and year to the proportion of the patient population with each strata for the corresponding state and year. All analyses were conducted using post-stratification weighted case-patient and patient-year counts to make nationally-representative inferences.

The calculation of post-stratification weights is shown in Equation 1, where *US*_*a*,*g*,*r*,*s*,*y*_ is the national population count in each age, gender, race and ethnicity, state, and year strata, according to data from the American Community Survey 5-year estimates;^1^ *US*_*s*,*y*_ is the annual state population; *p*_*a*,*g*,*r*,*s*,*y*_ is the total number of OERWD patients in each age, gender, race and ethnicity, state, and year strata; and *p*_*s*,*y*_ is number of OERWD patients per state and year (Equation 1).

**Equation 1. Calculation of post-stratification weights for Oracle EHR Real World Data (OERWD)**

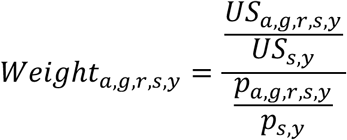

Note: *a* = age group, *g* = gender, *r* = race and ethnicity, *s* = state, *y* = year, *US* = American Community Survey population, *p* = patient population

#### Direct standardization

Our method for direct standardization is shown as in Equation 2a (for overall sIRs by climate region) and Equation 2b (for year-specific sIRs by climate region), where *x*_*c*,*a*,*g*,*r*_is the number of cases among a specific climate region (*c*), age group (*a*), gender (*g*), and race and ethnicity (*r*) weighted by selection weights (from Equation 1); *z*_*c*,*a*,*g*,*r*_is the strata-specific number of OERWD patient-years weighted by selection weights (from Equation 1); *t*_*a*,*g*,*r*_is the age-, gender-, and race and ethnicity-specific person-years among the total US patient population (i.e., standardizing population); *t* is the total patient-years among the US population; *n*_*a*_, *n*_*g*_, and *n*_*r*_ represent the total number of age, gender, and race and ethnicity strata, respectively; and *y* indicates year.

**Equation 2a. Age-, gender-, and race and ethnicity-standardized incidence rate for the study period**

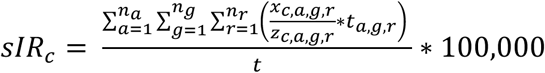

**Equation 2b. Age-, gender-, and race andethnicity-standardized incidence rate by year**

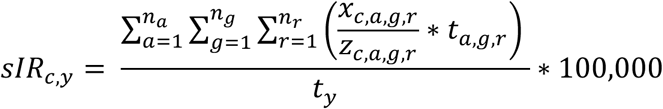

Note: *c* = climate region, *a* = age group, *g* = gender, *r* = race and ethnicity, *s* = state; *n*_*a*_, *n*_*g*_, and *n*_*r*_ represent the total number of age, gender, and race and ethnicity strata, respectively; *y* = year, *t* = total patient-years, *z* = patient population, *x* = number of case-patients

#### Standardized incidence rate ratios

To evaluate whether specific climate regions, age groups, genders, and racial and ethnic groups had disproportionately high incidence of blastomycosis or histoplasmosis, we computed overall and year-specific standardized incidence rate ratios (sIRRs) (Equations 3a and 3b). We calculated sIRRs for each region or group by dividing the region- or group-specific sIR by the overall incidence rates in the standard (national) population. For example, to compute sIRRs for each climate region, we calculated the age-, gender-, and race and ethnicity-standardized incidence rate per climate region and divided it by the incidence rate in the standard national population.

**Equation 3a. Standardized incidence rate ratio for the study period**

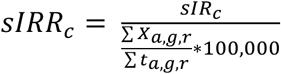

**Equation 3b. Annual standardized incidence rate ratio**

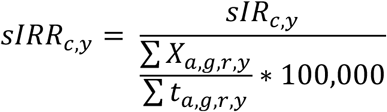

Note: *c* = climate region, *a* = age group, *g* = gender, *r* = race and ethnicity, *y* = year, *t* = total patient-years, *X* = number of case-patients

To estimate differences in incidence by racial and ethnic groups, we first age-, gender- and state-standardized incidence rates (sIRs) using the direct method and 5-year age groups based on the midpoint of annual population estimates of 5-year ACS data (Equation 4), where *x*_*r*,*a*,*g*,*s*_ is the number of cases among a specific race and ethnicity (*r*), gender (*g*), and state (*s*); *z*_*r*,*a*,*g*,*s*_ is the strata-specific number of OERWD patient-years; *t*_*a*,*g*,*s*_is the age-, gender-, and state-specific patient-years among the total US patient population (i.e., standardizing population); *t* is the total patient-years among the US population; *n*_*a*_, *n*_*g*_, and *n*_*s*_ represent the total number of age, gender, and state strata, respectively. We then computed sIRRs for each racial group and gender as described above using Equation 3a, exchanging race (Equation 4a), and gender (Equation 4b) for state as a strata. 95% confidence intervals for sIRs and sIRRs were calculated in the same manner as described above. To examine whether age groups or genders had disproportionately high incidence rates compared to their national representation, sIRs and sIRRs for age group and gender were also calculated with 95% CIs.

**Equation 4a. Age-, gender-, and state-standardized incidence rate for the study period by racial and ethnic group**

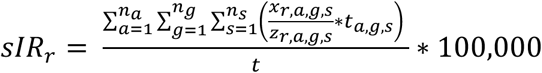

Note: *a* = age group, *g* = gender, *r* = race and ethnicity, *s* = state; *n*_*a*_, *n*_*g*_, and *n*_*s*_ represent the total number of age, gender, and state strata, respectively; *t* = total patient-years, *z* = patient population, *x* = number of case-patients

**Equation 4b. Age-, race and ethnicity-, and state-standardized incidence rate for the study period by gender**

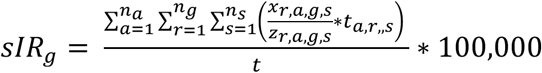

Note: *a* = age group, *g* = gender, *r* = race and ethnicity, *s* = state; *n*_*a*_, *n*_*g*_, and *n*_*s*_ represent the total number of age, race and ethnicity, and state strata, respectively; *t* = total patient-years, *z* = patient population, *x* = number of case-patients

## Supplemental Tables

**Table S1.** Incidence rate of blastomycosis and histoplasmosis per 100,000 patient-years by state and time period, United States, 2013-2023.

| State | Climate Region | Incidence rate per 100,000 patient-years |  |  |  |  |  |  |  |  |  |
| --- | --- | --- | --- | --- | --- | --- | --- | --- | --- | --- | --- |
|  |  | Blastomycosis |  |  |  |  | Histoplasmosis |  |  |  |  |
|  |  | 2013-2023 | 2013-2015 | 2016-2018 | 2019-2021 | 2022-2023 | 2013-2023 | 2013-2015 | 2016-2018 | 2019-2021 | 2022-2023 |
| Alabama | Southeast | 0.4 | 0.0 | 0.0 | 0.4 | 0.8 | 3.6 | 1.8 | 2.7 | 3.0 | 5.8 |
| Alaska | NA | 0.0 | 0.0 | 0.0 | 0.0 | 0.0 | 1.0 | 0.0 | 3.8 | 0.0 | 0.0 |
| Arizona | Southwest | 0.2 | 0.3 | 0.3 | 0.1 | 0.2 | 1.7 | 2.0 | 1.9 | 1.7 | 1.2 |
| Arkansas | South | 2.6 | 0.0 | 0.0 | 0.0 | 10.7 | 11.0 | 10.4 | 11.1 | 9.2 | 13.4 |
| California | West | 0.9 | 0.2 | 1.0 | 0.9 | 1.2 | 1.7 | 1.1 | 2.1 | 1.8 | 1.6 |
| Colorado | Southwest | 0.2 | 0.2 | 0.1 | 0.2 | 0.0 | 1.2 | 0.9 | 1.6 | 0.9 | 1.5 |
| Connecticut | Northeast | 0.2 | 0.2 | 0.4 | 0.1 | 0.0 | 0.7 | 0.8 | 0.6 | 0.6 | 0.8 |
| Delaware | Northeast | 0.2 | 0.0 | 0.2 | 0.0 | 0.4 | 1.8 | 0.2 | 2.4 | 2.1 | 1.5 |
| District of Columbia | Northeast | 0.3 | 0.0 | 1.1 | 0.0 | 0.0 | 1.8 | 0.0 | 0.3 | 2.1 | 5.9 |
| Florida | Southeast | 0.2 | 0.1 | 0.2 | 0.3 | 0.1 | 1.3 | 0.9 | 1.7 | 1.3 | 1.0 |
| Georgia | Southeast | 0.6 | 0.0 | 1.2 | 0.9 | 0.0 | 4.6 | 4.9 | 5.0 | 4.3 | 4.4 |
| Hawaii | NA | 0.5 | 0.0 | 0.9 | 0.5 | 0.7 | 0.2 | 0.0 | 0.0 | 0.3 | 0.5 |
| Idaho | Northwest | 0.4 | 0.0 | 0.0 | 1.0 | 0.0 | 0.2 | 0.0 | 0.0 | 0.0 | 0.8 |
| Illinois | Ohio Valley | 2.6 | 1.8 | 2.4 | 2.2 | 4.1 | 8.2 | 5.6 | 8.4 | 7.9 | 10.5 |
| Indiana | Ohio Valley | 1.8 | 1.7 | 1.4 | 2.0 | 1.9 | 16.8 | 10.4 | 14.6 | 17.0 | 21.0 |
| Iowa | Upper Midwest | 0.5 | 0.0 | 0.6 | 0.6 | 0.9 | 14.8 | 5.0 | 14.7 | 17.3 | 20.7 |
| Kansas | South | 0.3 | 0.0 | 0.1 | 0.3 | 0.6 | 21.9 | 6.1 | 18.4 | 22.3 | 28.6 |
| Kentucky | Ohio Valley | 1.6 | 2.0 | 1.7 | 0.3 | 2.0 | 16.3 | 17.1 | 21.1 | 11.8 | 11.9 |
| Louisiana | South | 0.0 | 0.0 | 0.0 | 0.0 | 0.0 | 0.0 | 0.0 | 0.0 | 0.0 | 0.0 |
| Maine | Northeast | 1.1 | 0.7 | 1.4 | 1.5 | 0.5 | 1.4 | 1.0 | 1.6 | 1.5 | 1.3 |
| Maryland | Northeast | 0.3 | 0.0 | 0.0 | 0.7 | 0.2 | 1.4 | 0.7 | 2.6 | 1.2 | 0.2 |
| Massachusetts | Northeast | 0.1 | 0.1 | 0.0 | 0.2 | 0.2 | 0.6 | 0.7 | 0.9 | 0.4 | 0.2 |
| Michigan | Upper Midwest | 12.3 | 0.0 | 0.0 | 27.1 | 6.2 | 8.0 | 16.7 | 5.4 | 5.6 | 9.0 |
| Minnesota | Upper Midwest | 2.5 | 1.4 | 1.5 | 3.8 | 3.5 | 4.9 | 3.3 | 4.4 | 7.4 | 4.4 |
| Mississippi | South | 0.9 | 0.0 | 0.9 | 0.6 | 1.5 | 2.4 | 3.6 | 0.6 | 3.0 | 3.4 |
| Missouri | Ohio Valley | 0.7 | 0.7 | 0.8 | 0.7 | 0.8 | 9.7 | 6.8 | 10.1 | 9.4 | 11.9 |
| Montana | N. Rockies | 0.4 | 0.0 | 0.7 | 0.3 | 0.4 | 1.0 | 0.8 | 1.0 | 0.9 | 1.4 |
| Nebraska | N. Rockies | 0.7 | 0.0 | 0.3 | 0.9 | 1.4 | 15.5 | 7.5 | 14.6 | 15.9 | 22.4 |
| Nevada | West | 2.1 | 0.0 | 6.5 | 0.9 | 0.0 | 1.3 | 0.0 | 0.5 | 0.0 | 5.4 |
| New Hampshire | Northeast | 0.3 | 0.0 | 0.0 | 0.2 | 0.6 | 1.0 | 0.0 | 0.8 | 0.8 | 1.5 |
| New Jersey | Northeast | 0.6 | 0.2 | 0.1 | 0.6 | 1.5 | 0.7 | 0.6 | 1.1 | 0.6 | 0.5 |
| New Mexico | Southwest | 0.5 | 0.0 | 0.3 | 0.9 | 0.5 | 2.2 | 0.6 | 2.9 | 1.9 | 3.0 |
| New York | Northeast | 1.1 | 0.4 | 1.3 | 1.7 | 0.9 | 1.1 | 1.1 | 1.2 | 0.9 | 1.4 |
| North Carolina | Southeast | 0.6 | 0.7 | 0.3 | 1.4 | 0.0 | 1.9 | 1.9 | 1.2 | 3.7 | 0.5 |
| North Dakota | N. Rockies | 1.2 | 0.0 | 0.0 | 4.2 | 0.0 | 0.0 | 0.0 | 0.0 | 0.0 | 0.0 |
| Ohio | Ohio Valley | 0.3 | 0.3 | 0.1 | 0.4 | 0.6 | 5.8 | 4.6 | 5.8 | 6.4 | 5.5 |
| Oklahoma | South | 0.0 | 0.0 | 0.0 | 0.0 | 0.0 | 12.0 | 10.4 | 11.2 | 9.9 | 17.6 |
| Oregon | Northwest | 0.2 | 0.4 | 0.1 | 0.3 | 0.2 | 1.1 | 0.6 | 0.9 | 2.2 | 0.5 |
| Pennsylvania | Northeast | 0.5 | 0.3 | 0.2 | 0.5 | 0.8 | 2.7 | 1.6 | 3.6 | 2.7 | 2.7 |
| Rhode Island | Northeast | 0.0 | 0.0 | 0.0 | 0.0 | 0.0 | 0.0 | 0.0 | 0.0 | 0.0 | 0.0 |
| South Carolina | Southeast | 0.2 | 0.0 | 0.1 | 0.3 | 0.6 | 2.1 | 1.7 | 2.6 | 1.6 | 2.5 |
| South Dakota | N. Rockies | 2.0 | 0.0 | 0.0 | 6.3 | 0.0 | 3.0 | 3.0 | 0.0 | 6.2 | 1.8 |
| Tennessee | Ohio Valley | 2.9 | 0.0 | 4.0 | 2.4 | 3.3 | 7.4 | 6.5 | 4.9 | 6.7 | 9.2 |
| Texas | South | 0.4 | 0.4 | 0.2 | 0.5 | 0.6 | 2.9 | 3.6 | 3.3 | 2.2 | 2.6 |
| Utah | Southwest | 0.2 | 0.0 | 0.0 | 0.7 | 0.0 | 0.9 | 0.8 | 0.5 | 1.5 | 0.6 |
| Vermont | Northeast | 1.5 | 1.3 | 2.5 | 0.5 | 1.8 | 1.5 | 2.3 | 1.8 | 1.1 | 0.7 |
| Virginia | Southeast | 0.2 | 0.0 | 0.1 | 0.5 | 0.0 | 2.2 | 0.1 | 3.3 | 2.9 | 1.3 |
| Washington | Northwest | 0.2 | 0.2 | 0.0 | 0.2 | 0.3 | 2.8 | 1.6 | 2.1 | 3.1 | 5.2 |
| West Virginia | Ohio Valley | 2.7 | 1.2 | 1.6 | 2.3 | 4.9 | 6.8 | 6.6 | 4.3 | 7.2 | 8.7 |
| Wisconsin | Upper Midwest | 7.4 | 2.1 | 6.9 | 7.4 | 9.9 | 5.2 | 4.1 | 5.6 | 5.2 | 5.5 |
| Wyoming | N. Rockies | 0.1 | 0.0 | 0.0 | 0.0 | 0.2 | 1.5 | 1.7 | 0.6 | 0.4 | 2.9 |

Note: N. Rockies = Northern Rockies and Plains. NOAA climate regions do not include Alaska or Hawaii.

**Table S2.** Incidence rate of blastomycosis and histoplasmosis per 100,000 patient-years by climate region, United States, 2013-2023.

|  |  | Incidence rate per 100,000 patient-years |  |
| --- | --- | --- | --- |
| Climate Region | Year | Blastomycosis | Histoplasmosis |
| All | 2013 | 0.4 | 2.9 |
|  | 2014 | 0.4 | 2.8 |
|  | 2015 | 0.5 | 3.4 |
|  | 2016 | 0.7 | 4.7 |
|  | 2017 | 0.7 | 5.0 |
|  | 2018 | 0.8 | 4.3 |
|  | 2019 | 0.8 | 5.1 |
|  | 2020 | 0.9 | 4.1 |
|  | 2021 | 1.1 | 5.2 |
|  | 2022 | 1.1 | 5.1 |
|  | 2023 | 1.3 | 6.5 |
| Northeast | 2013 | 0.2 | 0.3 |
|  | 2014 | 0.4 | 1.0 |
|  | 2015 | 0.2 | 1.4 |
|  | 2016 | 0.3 | 2.2 |
|  | 2017 | 0.7 | 2.0 |
|  | 2018 | 0.3 | 1.5 |
|  | 2019 | 0.4 | 1.3 |
|  | 2020 | 0.8 | 1.1 |
|  | 2021 | 0.7 | 1.6 |
|  | 2022 | 0.6 | 1.5 |
|  | 2023 | 0.7 | 1.4 |
| Northern Rockies and Plains | 2013 | 0 | 1.8 |
|  | 2014 | 0 | 2.5 |
|  | 2015 | 0 | 7.0 |
|  | 2016 | 0 | 4.8 |
|  | 2017 | 0.6 | 9.0 |
|  | 2018 | 0.7 | 7.4 |
|  | 2019 | 0.3 | 6.2 |
|  | 2020 | 0.7 | 8.4 |
|  | 2021 | 1.1 | 8.3 |
|  | 2022 | 0.9 | 11.2 |
|  | 2023 | 0.7 | 11.0 |
| Northwest | 2013 | 0.7 | 0.3 |
|  | 2014 | 0.0 | 0.8 |
|  | 2015 | 0.4 | 1.2 |
|  | 2016 | 0.0 | 1.5 |
|  | 2017 | 0.0 | 0.8 |
|  | 2018 | 0.3 | 1.4 |
|  | 2019 | 0.2 | 1.7 |
|  | 2020 | 0.5 | 0.7 |
|  | 2021 | 0.3 | 4.0 |
|  | 2022 | 0.2 | 0.7 |
|  | 2023 | 0.3 | 3.1 |
| Ohio Valley | 2013 | 1.1 | 9.5 |
|  | 2014 | 1.3 | 8.5 |
|  | 2015 | 1.3 | 8.4 |
|  | 2016 | 1.4 | 10.8 |
|  | 2017 | 1.3 | 12.2 |
|  | 2018 | 1.3 | 9.5 |
|  | 2019 | 1.3 | 11.0 |
|  | 2020 | 1.4 | 9.5 |
|  | 2021 | 1.8 | 12.2 |
|  | 2022 | 1.9 | 12.2 |
|  | 2023 | 2.3 | 15.5 |
| South | 2013 | 0.5 | 6.0 |
|  | 2014 | 0.3 | 1.9 |
|  | 2015 | 0.3 | 4.1 |
|  | 2016 | 0.0 | 7.5 |
|  | 2017 | 0.3 | 5.3 |
|  | 2018 | 0.3 | 7.1 |
|  | 2019 | 0.5 | 10.6 |
|  | 2020 | 0.5 | 6.4 |
|  | 2021 | 0.3 | 6.0 |
|  | 2022 | 0.4 | 7.2 |
|  | 2023 | 1.1 | 13.3 |
| Southeast | 2013 | 0.3 | 1.4 |
|  | 2014 | 0.0 | 1.7 |
|  | 2015 | 0.0 | 1.2 |
|  | 2016 | 0.2 | 1.8 |
|  | 2017 | 0.3 | 2.9 |
|  | 2018 | 0.2 | 2.5 |
|  | 2019 | 0.5 | 3.3 |
|  | 2020 | 0.5 | 1.4 |
|  | 2021 | 0.4 | 1.8 |
|  | 2022 | 0.2 | 1.5 |
|  | 2023 | 0.2 | 2.1 |
| Southwest | 2013 | 0.3 | 2.0 |
|  | 2014 | 0.0 | 1.6 |
|  | 2015 | 0.4 | 1.3 |
|  | 2016 | 0.3 | 2.2 |
|  | 2017 | 0.3 | 1.6 |
|  | 2018 | 0.3 | 2.1 |
|  | 2019 | 0.3 | 2.2 |
|  | 2020 | 0.2 | 1.4 |
|  | 2021 | 0.2 | 1.3 |
|  | 2022 | 0.1 | 1.3 |
|  | 2023 | 0.2 | 1.6 |
| Upper Midwest | 2013 | 0.5 | 2.8 |
|  | 2014 | 0.3 | 4.7 |
|  | 2015 | 1.8 | 5.6 |
|  | 2016 | 1.7 | 8.2 |
|  | 2017 | 1.5 | 11.8 |
|  | 2018 | 4.2 | 9.0 |
|  | 2019 | 3.4 | 11.0 |
|  | 2020 | 3.7 | 7.4 |
|  | 2021 | 6.5 | 12.0 |
|  | 2022 | 4.7 | 10.7 |
|  | 2023 | 6.8 | 10.4 |
| West | 2013 | 0.4 | 0.4 |
|  | 2014 | 0.1 | 0.7 |
|  | 2015 | 0.2 | 1.8 |
|  | 2016 | 0.9 | 2.7 |
|  | 2017 | 0.7 | 1.9 |
|  | 2018 | 1.5 | 1.6 |
|  | 2019 | 1.2 | 1.9 |
|  | 2020 | 0.9 | 1.4 |
|  | 2021 | 0.7 | 2.0 |
|  | 2022 | 1.2 | 1.4 |
|  | 2023 | 1.2 | 1.9 |

Note: NOAA climate regions do not include Alaska or Hawaii.

**Table S3.**
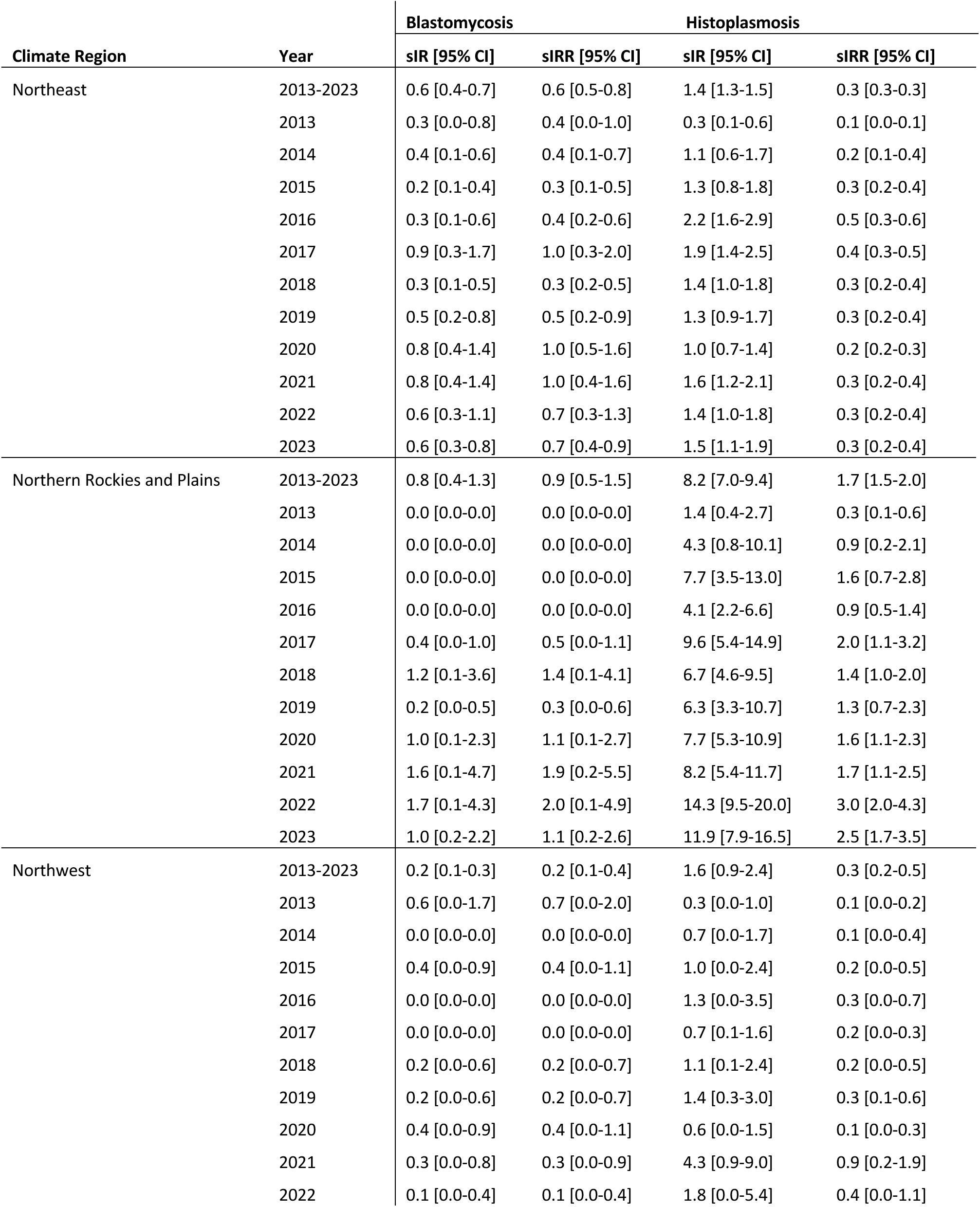
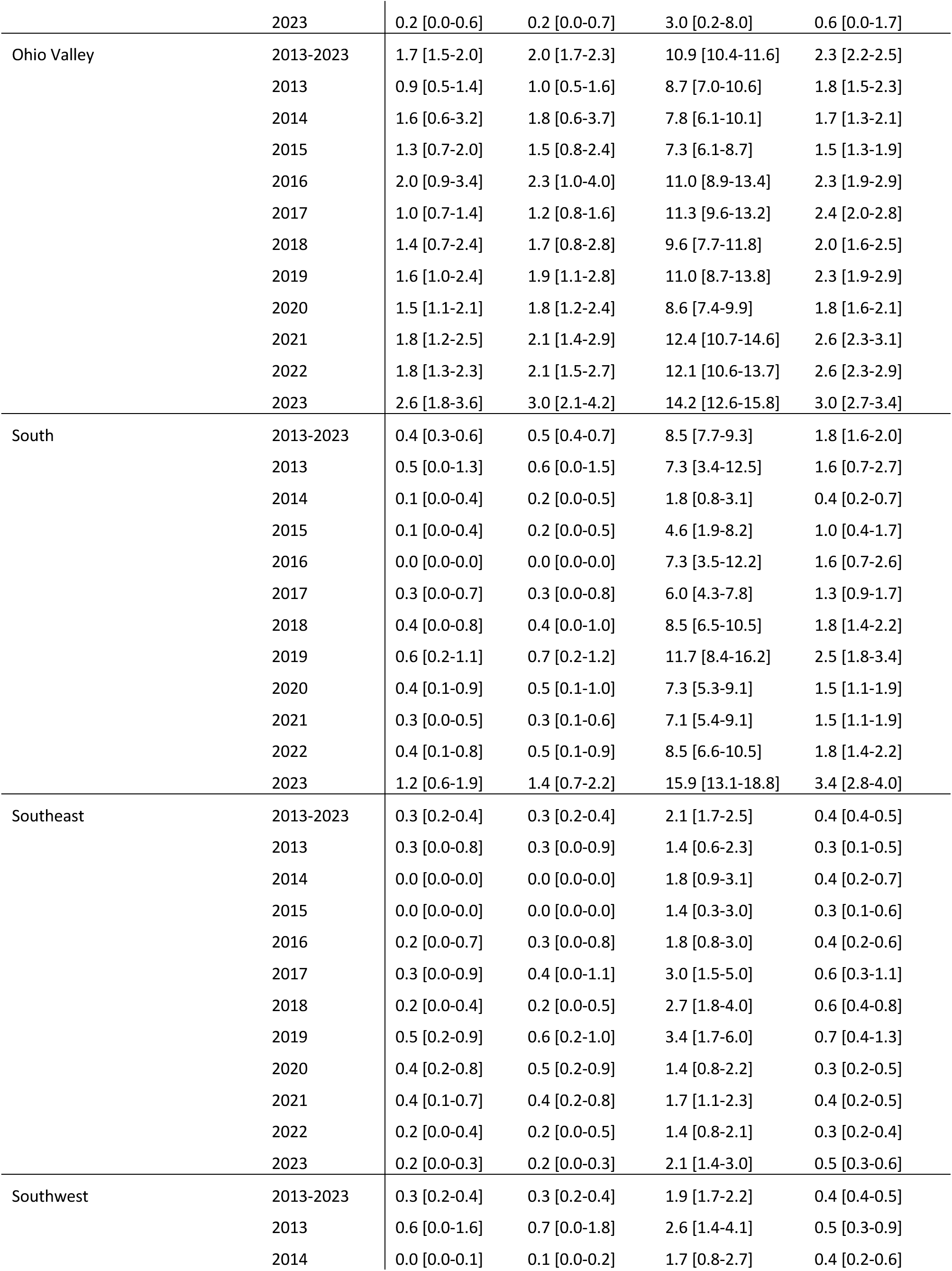
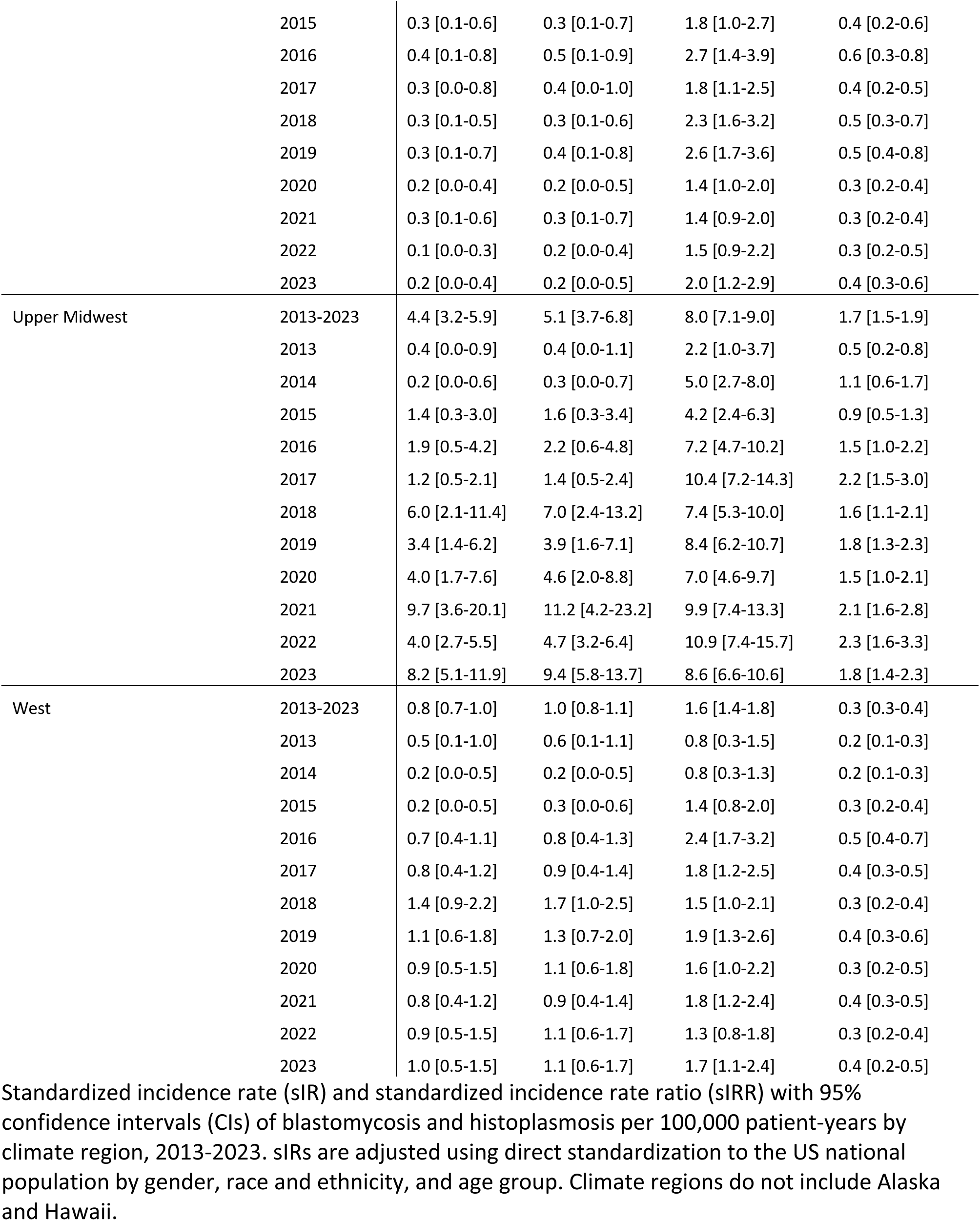
Standardized incidence rates and standardized incidence rate ratios of blastomycosis and histoplasmosis per 100,000 patient-years by climate region and year, United States, 2013-2023.

**Table S4.** Standardized incidence rate and standardized incidence rate ratio of blastomycosis and histoplasmosis per 100,000 patient-years by age group, race and ethnicity, and gender, United States, 2013-2023.

| Age Group | Blastomycosis |  | Histoplasmosis |  |
| --- | --- | --- | --- | --- |
|  | sIR [95% CI] | sIRR [95% CI] | sIR [95% CI] | sIRR [95% CI] |
| <5 | 0.1 [0.0-0.1] | 0.1 [0.0-0.1] | 0.3 [0.2-0.4] | 0.1 [0.0-0.1] |
| 5-9 | 0.2 [0.1-0.2] | 0.2 [0.1-0.3] | 0.7 [0.6-0.9] | 0.2 [0.1-0.2] |
| 10-14 | 0.2 [0.1-0.3] | 0.2 [0.1-0.3] | 1.5 [1.3-1.8] | 0.3 [0.3-0.4] |
| 15-19 | 0.4 [0.3-0.6] | 0.5 [0.4-0.7] | 2.9 [2.5-3.3] | 0.6 [0.5-0.7] |
| 20-24 | 0.4 [0.2-0.5] | 0.4 [0.3-0.6] | 2.3 [1.9-2.7] | 0.5 [0.4-0.6] |
| 25-29 | 0.6 [0.4-0.8] | 0.7 [0.5-0.9] | 2.9 [2.5-3.4] | 0.6 [0.5-0.7] |
| 30-34 | 0.8 [0.6-1.0] | 0.9 [0.6-1.1] | 3.5 [3.1-4.0] | 0.7 [0.7-0.8] |
| 35-39 | 0.6 [0.5-0.8] | 0.7 [0.5-1.0] | 4.4 [3.9-4.9] | 0.9 [0.8-1.0] |
| 40-44 | 0.8 [0.6-1.0] | 0.9 [0.6-1.1] | 5.1 [4.6-5.7] | 1.1 [1.0-1.2] |
| 45-49 | 0.9 [0.7-1.2] | 1.1 [0.8-1.4] | 6.4 [5.8-7.0] | 1.4 [1.2-1.5] |
| 50-54 | 0.9 [0.7-1.1] | 1.0 [0.8-1.3] | 6.9 [6.2-7.8] | 1.5 [1.3-1.6] |
| 55-59 | 1.4 [1.0-1.7] | 1.6 [1.2-2.0] | 7.0 [6.4-7.6] | 1.5 [1.4-1.6] |
| 60-64 | 1.6 [1.3-1.9] | 1.8 [1.5-2.1] | 7.9 [7.1-8.9] | 1.7 [1.5-1.9] |
| 65+ | 1.9 [1.7-2.1] | 2.2 [1.9-2.4] | 8.4 [7.9-8.9] | 1.8 [1.7-1.9] |
| **Race and Ethnicity** |  |  |  |  |
| Hispanic or Latino | 1.4 [0.9-2.1] | 1.6 [1.0-2.4] | 3.8 [3.1-4.4] | 0.8 [0.7-0.9] |
| NH American Indian or Alaska Native | 0.9 [0.3-1.9] | 1.1 [0.3-2.2] | 3.2 [1.1-5.9] | 0.7 [0.2-1.3] |
| NH Asian | 1.2 [0.6-1.9] | 1.4 [0.7-2.1] | 2.7 [2.0-3.4] | 0.6 [0.4-0.7] |
| NH Black or African American | 1.1 [0.9-1.4] | 1.3 [1.0-1.6] | 4.2 [3.7-4.6] | 0.9 [0.8-1.0] |
| NH Other Race | 0.5 [0.3-0.7] | 0.5 [0.3-0.8] | 4.7 [3.1-6.9] | 1.0 [0.7-1.5] |
| NH White | 0.8 [0.7-0.8] | 0.9 [0.8-0.9] | 4.9 [4.8-5.1] | 1.0 [1.0-1.1] |
| **Gender** |  |  |  |  |
| Female | 0.4 [0.4-0.5] | 0.5 [0.4-0.6] | 4.1 [3.9-4.2] | 0.9 [0.8-0.9] |
| Male | 1.3 [1.2-1.4] | 1.5 [1.4-1.6] | 5.3 [5.1-5.6] | 1.1 [1.1-1.2] |

Note: Standardized incidence rate (sIR) and standardized incidence rate ratio (sIRR) with 95% confidence intervals of blastomycosis and histoplasmosis per 100,000 patient-years by age group (years), race and ethnicity, and gender, 2013-2023. sIRs are adjusted using direct standardization to the US national population for gender, race and ethnicity, and state (for age group sIRs); gender, state, and age group (for race and ethnicity sIRs); and age group, race and ethnicity, and state (for gender sIRs). NH = non-Hispanic.

## Footnotes

1 US Census Bureau. American Community Survey 5-Year Data (2009-2023). Available at: https://www.census.gov/data/developers/data-sets/acs-5year.html. Accessed 22 January 2025.

